# A workforce up in smoke? Examining trends in health-related economic inactivity by smoking status in England, 2013-2025

**DOI:** 10.1101/2025.05.05.25327007

**Authors:** Sarah Jackson, Sharon Cox, Jamie Brown

## Abstract

**Objectives:** To examine time trends in health-related economic inactivity among working-age adults in England by smoking status between 2013 and 2025, and to estimate the number of smokers economically inactive due to ill health in 2025.

**Design:** Repeat cross-sectional analysis of data from a nationally representative survey (the Smoking Toolkit Study).

**Setting:** England.

**Participants:** 173,248 adults aged 18-64y surveyed between March 2013 and February 2025.

**Main outcome measures:** Health-related economic inactivity was defined as not being in paid work due to long-term illness or disability. Logistic regression modelled time trends adjusted for age and gender, by (i) smoking status (current, former, never) and (ii) duration of abstinence among former smokers. National population and smoking prevalence data were used to estimate absolute numbers of inactive smokers.

**Results:** Across the period, health-related economic inactivity more than doubled in all adults (2.5% [2.3-2.7%] to 5.5% [5.1-5.9%]; prevalence ratio PR=2.21 [1.96-2.49]). Prevalence was consistently highest - and absolute increases over time were largest - among current smokers; reaching 11.3% [9.9-12.7%] in 2025, compared with 5.8% [5.0-6.6%] in former smokers and 3.3% [2.9-3.7%] in never smokers. This equates to ∼750,000 current smokers not in work due to ill health or disability in 2025, up from ∼390,000 in 2013. Among former smokers, inactivity was highest in recent quitters and declined with longer abstinence duration.

**Conclusions:** Current smoking is strongly associated with health-related economic inactivity, and this disparity has widened in absolute terms over time. In early 2025, one in nine working-age adults in England who smoked was not in work due to long-term illness or disability. Efforts to reduce smoking prevalence may contribute to tackling rising inactivity and improving labour market participation.

## Introduction

The UK is currently facing an employment crisis, with a significant number of working-age individuals out of the labour market due to ill health or disability. According to the Office for National Statistics (ONS), a quarter of the working-age population (16-64y) do not currently have a job,^1^ of whom approximately 2.7 million are not in work due to ill health.^2^ Around half of people with disabilities are not in paid work – a rate more than double that of the rest of the working-age population.^2^ Economic inactivity (defined as not actively looking for work or not available to start a job) appears to have increased since the COVID-19 pandemic^1,3^ – driven largely by long-term physical and mental ill health and disability^4^ – placing a strain on public services, reducing productivity, and contributing to stalls in economic growth. However, there has been some debate about the extent of the rise,^5,6^ with an independent report suggesting it may be partly due to bias in who has responded to the ONS’ labour force survey (the main source of data on unemployment).^5^ Addressing the causes of work incapacity is a key government priority, as reflected in policies aimed at reducing long-term sickness-related unemployment and boosting workforce participation.^7^

It is well established that smoking-related ill health contributes to economic inactivity.^8–11^ Smoking is a modifiable behaviour with severe disease outcomes (e.g., chronic obstructive pulmonary disease, cancers, and cardiovascular disease) that lead to and exacerbate disabilities, all of which can result in long-term work absence and economic disengagement.^12–14^ Smoking is also associated with poor mental health,^15–17^ which is thought to have been an important driver of recent increases in long-term sickness absence.^3,18^ Severe psychological distress has become more prevalent in Great Britain since the pandemic, with particularly high levels among people who smoke.^19^ Estimates suggest smoking costs the economy in England in the region of £27.6 billion each year in lost productivity due to smoking-related unemployment, lost earnings, and premature mortality.^11,20^

Understanding the current importance of smoking in health-related economic inactivity can provide insights into how reducing smoking rates might benefit society more broadly, such as improving employment rates. Using data from a nationally-representative household survey in England conducted between 2013 and 2025, this study aimed to examine time trends in the age- and gender-adjusted proportion of adults not in work due to long-term illness or disability, among all working-age adults and by (i) smoking status (current, former, or never smoker) and (ii) among former smokers, duration of abstinence. We used national estimates of population size and smoking prevalence to estimate the number of adult smokers in England currently not in work due to long-term illness or disability. In addition, we characterised the recent sociodemographic profile of this group and compared it to never and former smokers currently not in work due to long-term illness or disability.

## Methods

### Pre-registration

The study protocol and analysis plan were pre-registered on Open Science Framework (https://osf.io/nygzr/). In addition to our pre-registered analyses, we included descriptive data on psychological distress among those not in work due to long-term illness or disability.

### Design

Data were drawn from the Smoking Toolkit Study (STS), an ongoing monthly cross-sectional survey of a representative sample of adults (≥16 years) in England.^21,22^ The STS uses a hybrid of random probability and simple quota sampling to select a new sample of approximately 1,700 adults each month. Data were collected face-to-face up to the start of the Covid-19 pandemic and via telephone from April 2020 onwards; the two modes show good comparability on key sociodemographic and smoking indices.^23^ Comparisons with other national surveys and sales data indicate the survey achieves nationally representative estimates of key variables such as sociodemographic characteristics, smoking prevalence, and cigarette consumption.^21,24^

The present analyses used data from respondents between March 2013 (the first wave to assess working status) and February 2025 (the most recent data at the time of analysis). Data were not collected from 16- and 17-year-olds between April 2020 and December 2021, so we restricted the sample to those aged ≥18y for consistency across the time series.

### Measures

Health-related economic inactivity was operationalised as participants reporting that they are ‘not in paid work because of long-term illness or disability’, in response to the following question assessing their current working status: ‘Which of these applies to you?’

1. Have paid job – full time (30+ hours per week)
2. Have paid job – part time (8-29 hours per week)
3. Have paid job – part time (under 8 hours per week)
4. Not working – housewife
5. Self-employed
6. Full time student
7. Still at school
8. Unemployed and seeking work
9. Retired
10. Not in paid work for other reason
11. Not in paid work because of long term illness or disability
12. Refused

Smoking status was assessed by asking participants which of the following best applied to them:

a. I smoke cigarettes (including hand-rolled) every day
b. I smoke cigarettes (including hand-rolled), but not every day
c. I do not smoke cigarettes at all, but I do smoke tobacco of some kind (e.g. pipe, cigar or shisha)
d. I have stopped smoking completely in the last year
e. I stopped smoking completely more than a year ago
f. I have never been a smoker (i.e. smoked for a year or more)

Those who responded a-c were considered current smokers. Those who responded d-e were considered former smokers. Those who responded f were considered never-smokers.

For former smokers, we calculated the duration of abstinence in years (i.e., how many years ago a participant quit smoking). For those who quit in the past year (response d), this was assessed with the question: ‘How long ago did your most recent serious quit attempt start? By most recent, we mean the last time you tried to quit’ with response options coded as follows:

1. In the last week [0.02 years]
2. More than a week and up to a month [0.05 years]
3. More than 1 month and up to 2 months [0.125 years]
4. More than 2 months and up to 3 months [0.21 years]
5. More than 3 months and up to 6 months [0.375 years]
6. More than 6 months and up to a year [0.75 years]

For those who quit more than a year ago (response e), it was calculated as the participant’s actual age minus the age when they stopped smoking. Duration of abstinence was analysed as a continuous variable (see *analyses*).

Age was analysed as a continuous covariate for trend analyses and categorised as 18-24, 25-34, 35-44, 45-54, 55-64, and ≥65 years for descriptive and age-stratified analyses. Gender was self-identified as man, woman, or in another way; we excluded the latter category for trend analyses due to the small number of participants but retained it for descriptive analyses. Region in England was categorised as North West, North East, Yorkshire and the Humber, West Midlands, East Midlands, East of England, South West, South East, and London.

Psychological distress was assessed using the Kessler Psychological Distress Scale (K6), which measures non-specific psychological distress in the past month (possible range 0-24);^25,26^ we coded scores ≤4 as no or low distress vs. 5-12 as moderate and ≥13 as severe distress.^25,27^ Between 2022 and 2025, this variable was only included in selected waves (January 2022-June 2023, January-March 2024, and January-February 2025), so analyses of distress are restricted to participants surveyed in these months.

### Statistical analysis

Data were analysed using R v.4.4.2. The STS uses raking to weight the sample to match the population in England. This profile is determined each month by combining data from the UK Census, the Office for National Statistics mid-year estimates, and the annual National Readership Survey.^21^ The following analyses used weighted data. We excluded participants who did not report their smoking or working status.

We used logistic regression to model time trends in the proportion of adults not in work due to long-term illness or disability between March 2013 and February 2025. To estimate changes over time among all working-age adults, we ran a model with time as the predictor. To examine differences by smoking status, we modelled the interaction between time and smoking status – thus allowing for time trends to differ between never, former, and current smokers. To examine differences among former smokers by duration of abstinence, we modelled the interaction between time and duration of abstinence. In an unplanned analysis, we repeated these models stratified by age to explore differences in time trends by age group.

For all models, time (survey wave) was modelled using restricted cubic splines, to allow for flexible and non-linear changes over time while avoiding categorisation. We compared models with three, four, and five knots using the Akaike Information Criterion (AIC) and reported the best-fitting model for each outcome (selected as the model with the lowest AIC value or the simplest model within 2 AIC units; **Table S1**). Duration of abstinence was modelled using restricted cubic splines, with three knots (placed at the 5^th^, 50^th^, and 95^th^ percentiles; sufficient to model non-linear associations while avoiding overfitting). All models were adjusted for age and gender.

To illustrate the extent of changes from the start to the end of the period within each subgroup, we reported the absolute percentage point change and the relative change (prevalence ratio [PR]) between March 2013 and February 2025, alongside 95% confidence intervals (CIs) calculated using bootstrapping (1,000 replications). We used predicted modelled estimates to plot the age- and gender-adjusted proportion of never, former, and current smokers not in work due to long-term illness or disability over the study period.

Using the Office for National Statistics’ (ONS) mid-year population estimates for England,^28^ we estimated the number of working-age current smokers not in work due to long-term illness or disability as of February 2025 (the final month of the study period). The calculation was as follows:

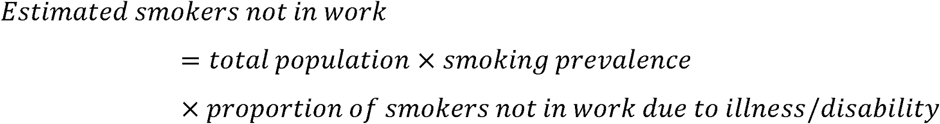

Where:

- Total population: Number of adults aged 18-64y in England (ONS 2023 mid-year estimates)
- Smoking prevalence: The proportion of working-age adults who currently smoke (STS; December 2024-February 2025 [we used data aggregated across the most recent three waves rather than just February 2025 to increase the sample size for this estimate])
- Proportion of smokers not in work due to illness/disability: Modelled estimate for February 2025 (STS)

For comparison, we also estimated the corresponding number of current smokers not in work due to long-term illness or disability in the first month of the study period (March 2013), using ONS 2013 mid-year estimates of population size, smoking prevalence in March-May 2013, and the modelled estimate for March 2013.

Finally, we used descriptive statistics to compare the profiles (i.e., sociodemographic characteristics) of current, former, and never smokers not in work due to long-term illness or disability. For this analysis, we restricted the sample to those surveyed between 2022 and 2025, to provide up-to-date information while ensuring sufficient sample sizes. Within each smoking status, we calculated the proportions (with 95% CIs) of those not in work due to long-term illness or disability belonging to each age group, gender, and region.

## Results

A total of 174,188 adults aged 18-64 years were surveyed in England between March 2013 and February 2025. We excluded 940 (0.5%) with missing data on working or smoking status, leaving a final sample of 173,248 participants, of whom 19.6% were current smokers, 18.2% were former smokers, and 62.2% had never regularly smoked. Sample characteristics, overall and by smoking status, are presented in **Table S2**.

### Time trends in health-related economic inactivity

Across the study period, the prevalence of health-related economic inactivity in all working-age adults more than doubled, rising from 2.5% [2.3-2.7%] in March 2013 to 5.5% [5.1-5.9%] in February 2025 (PR=2.21 [1.96-2.49]; **Table 1**). This increase was non-linear (**Figure 1**), with most of the growth occurring after the start of the pandemic (prevalence in March 2020: 3.2% [3.0-3.3%]; **Table S3**).

**Figure 1.**
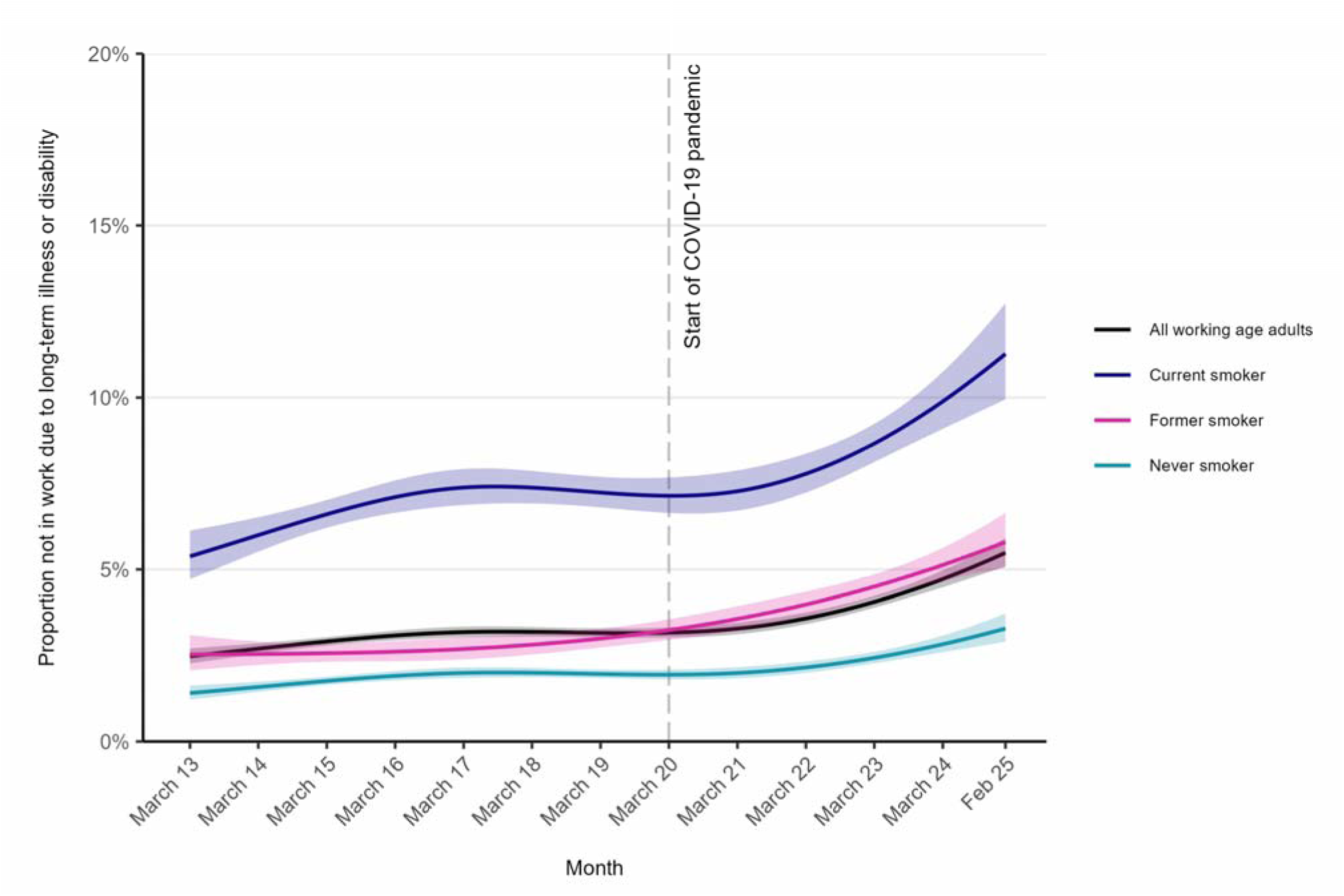
Trends in health-related economic inactivity among working-age adults in England, overall and by smoking status, March 2013 to February 2025. Lines represent modelled weighted prevalence by monthly survey wave, modelled non-linearly using restricted cubic splines (see **Table S1** for model selection), adjusted for age and gender. Shaded bands represent 95% confidence intervals.

**Table 1.** Modelled estimates of changes in the proportion of working-age adults in England not in work due to long-term illness or disability.

|  | % [95%CI] not in work due to long-term illness or disability<sup>1</sup> |  | Absolute percentage point change [95% CI]<sup>2</sup> | Relative change, prevalence ratio [95% CI]<sup>3</sup> |
| --- | --- | --- | --- | --- |
|  | March 2013 | February 2025 |  |  |
| All working-age adults | 2.5 [2.3–2.7] | 5.5 [5.1–5.9] | 3.0 [2.5–3.5] | 2.21 [1.96–2.49] |
| By smoking status |  |  |  |  |
| Never | 1.4 [1.2–1.6] | 3.3 [2.9–3.7] | 1.9 [1.4–2.4] | 2.33 [1.89–2.89] |
| Former | 2.5 [2.1–3.1] | 5.8 [5.0–6.6] | 3.3 [2.3–4.2] | 2.29 [1.78–2.98] |
| Current | 5.4 [4.7–6.1] | 11.3 [9.9–12.7] | 5.9 [4.2–7.5] | 2.10 [1.71–2.52] |
| By duration of abstinence, among former smokers<sup>4</sup> |  |  |  |  |
| 1 year | 4.7 [3.4–6.5] | 12.1 [9.6–15.1] | 7.4 [4.8–10.8] | 2.54 [1.85–3.82] |
| 5 years | 3.8 [3.0–4.7] | 9.2 [8.0–10.7] | 5.5 [4.2–7.1] | 2.45 [2.00–3.16] |
| 10 years | 3.0 [2.3–3.8] | 6.8 [5.7–8.0] | 3.8 [2.7–5.0] | 2.28 [1.82–3.03] |
| 20 years | 2.3 [1.7–3.1] | 4.6 [3.8–5.7] | 2.3 [1.4–3.4] | 2.01 [1.53–2.85] |

CI, confidence interval.
<sup>1</sup> Data are weighted estimates of prevalence in the first and last months in the study period, from logistic regression with survey month modelled non-linearly using restricted cubic splines (**Table S1** for model selection), adjusted for age and gender. Modelled estimates in each year are provided in **Table S3**.
<sup>2</sup> Absolute percentage point change calculated as prevalence in February 2025 minus prevalence in March 2013 with 95% CIs calculated using bootstrapping (1,000 replications).
<sup>3</sup> Prevalence ratio calculated as prevalence in February 2025 divided by prevalence in March 2013 with 95% CIs calculated using bootstrapping (1,000 replications).
<sup>4</sup> Modelled estimates are shown for selected durations of abstinence to illustrate differences. Note that the model used to derive these estimates included data from former smokers with any duration of abstinence, not only those abstinent for 1, 5, 10, or 20 years.

At all timepoints, the prevalence of health-related economic inactivity was highest among current smokers and lowest among never smokers (**Figure 1**). While relative increases in prevalence over time were broadly similar across smoking statuses, absolute increases were largest among current smokers (+5.9 percentage points [ppts]) compared with former (+3.3 ppts) and never smokers (+1.9 ppts; **Table 1**). By February 2025, 11.3% of working-age current smokers were not in work due to long-term illness or disability – nearly double the rate among former smokers (5.8%) and more than three times that of never smokers (3.3%).

When extrapolated to the national population, these figures suggest that approximately 750,000 current smokers were economically inactive due to ill health in early 2025 (34.9 million working-age adults x 19.1% current smoking prevalence x 11.3%), up from around 390,000 in 2013 (33.1 million x 21.8% x 5.4%). This is despite the overall decline in national smoking prevalence. The equivalent numbers for all working-age adults were 1.9 million in early 2025 (34.9 million x 5.5%), up from 830,000 in 2013 (33.1 million x 2.5%).

Exploratory analyses stratified by age group showed the prevalence of health-related economic inactivity was higher, and absolute increases over time were greater, with increasing age (**Figure S1**). For example, prevalence increased from 4.7% in March 2013 to 11.0% in February 2025 (+6.3 ppts) among those aged 55-64 compared with 0.7% to 1.8% (+1.1 ppts) among those aged 18-24 (**Table S4**). In addition, differences by smoking status were much more pronounced in older age groups (**Figure S1, Table S4**).

Among former smokers, the prevalence of health-related economic inactivity was consistently higher among those with shorter durations of abstinence (**Figure 2**). Absolute increases over time were largest among more recent quitters – for example, +7.4 ppts among those who quit one year ago compared with +2.3 ppts among those who quit 20 years ago – though relative increases were similar across durations of abstinence (**Table 1**).

**Figure 2.**
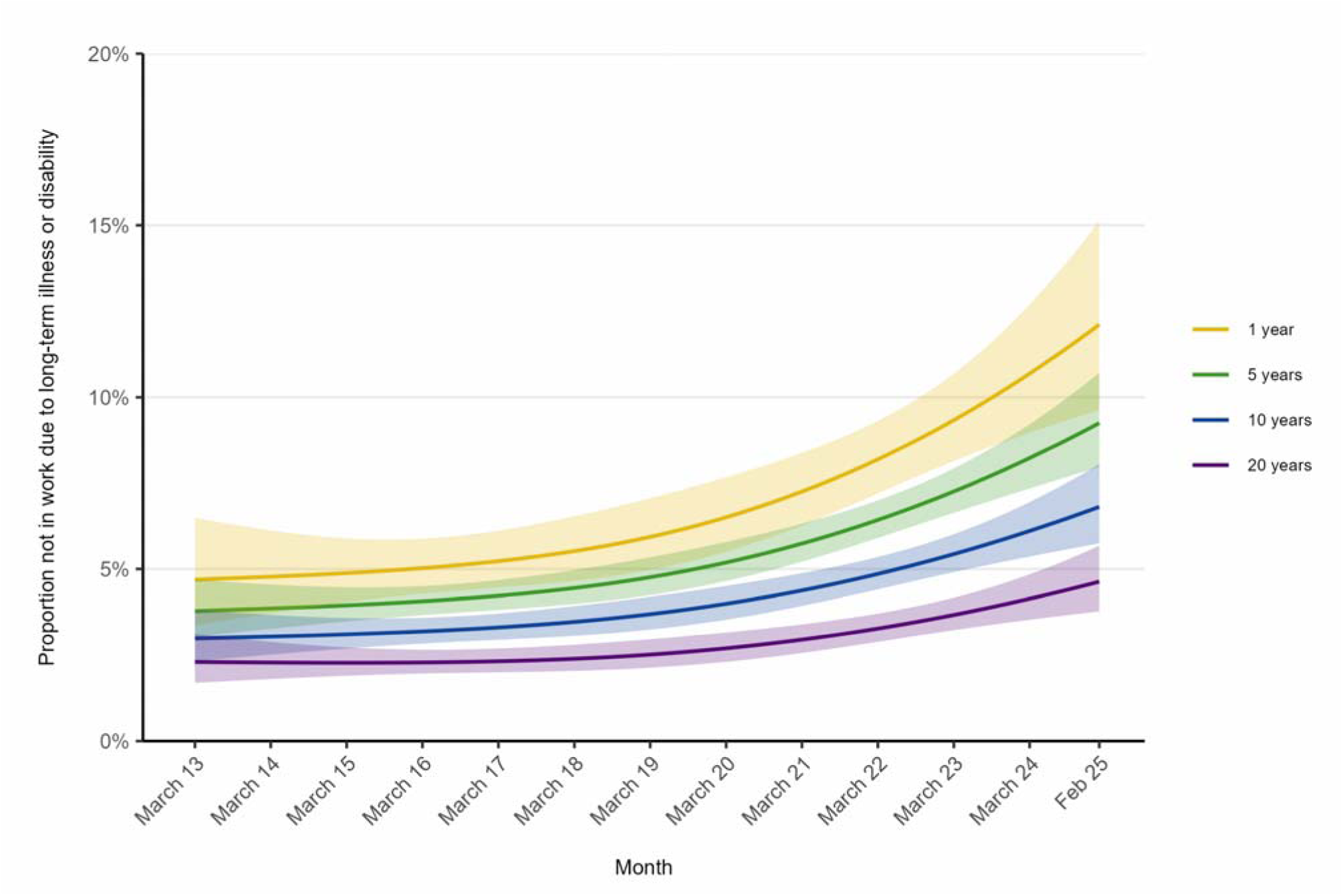
Trends in health-related economic inactivity among working-age former smokers in England by duration of abstinence, March 2013 to February 2025. Lines represent modelled weighted prevalence by monthly survey wave and duration of abstinence, both modelled non-linearly using restricted cubic splines (see **Table S1** for model selection), adjusted for age and gender. Shaded bands represent 95% confidence intervals. Modelled estimates are shown for selected durations of abstinence to illustrate differences. Note that the model used to derive these estimates included data from former smokers with any duration of abstinence, not only those abstinent for 1, 5, 10, or 20 years.

### Sociodemographic profile of those not in work due to long-term illness or disability

In 2022-25, current smokers who were not in work due to long-term illness or disability were on average 45.5 years old; one year younger than those who had never regularly smoked and three years younger than former smokers (**Table 2**). This pattern reflects broader age differences by smoking status (**Table S2**). A quarter (25.2%) of current smokers not in work due to long-term illness or disability were aged under 35. More than half of those not in work due to long-term illness or disability were women, including 56.2% of current smokers (**Table 2**). The proportion who described their gender in another way (i.e., not as a man or woman) was lowest among current smokers. There were no notable differences in the regional distribution of those not in work due to long-term illness or disability by smoking status (**Table 2**).

**Table 2.** Sociodemographic and psychological profile of working-age adults not in work due to long-term illness or disability in 2022-25, overall and by smoking status.

|  | Not in work due to long-term illness or disability |  |  |  |
| --- | --- | --- | --- | --- |
|  | All working-age adults | Never smokers | Former smokers | Current smokers |
| Unweighted *N* | 2,254 | 809 | 672 | 773 |
| Age (years) |  |  |  |  |
| Mean [95% CI] | 46.8 [46.2–47.3] | 46.5 [45.6–47.5] | 48.5 [47.5–49.5] | 45.5 [44.6–46.4] |
| 18-24 | 5.3 [4.2–6.3] | 5.8 [3.9–7.6] | 4.6 [2.8–6.3] | 5.3 [3.6–7.0] |
| 25-34 | 16.3 [14.6–18.0] | 15.9 [13.0–18.7] | 12.7 [9.8–15.6] | 19.9 [16.9–23.0] |
| 35-44 | 18.5 [16.8–20.3] | 19.3 [16.3–22.3] | 17.5 [14.3–20.7] | 18.6 [15.6–21.6] |
| 45-54 | 26.6 [24.7–28.6] | 26.4 [23.0–29.7] | 25.5 [22.0–29.0] | 27.9 [24.5–31.3] |
| 55-64 | 33.3 [31.2–35.3] | 32.7 [29.3–36.1] | 39.7 [35.8–43.6] | 28.3 [25.0–31.5] |
| Gender |  |  |  |  |
| Man | 39.5 [37.3–41.7] | 38.8 [35.1–42.4] | 36.7 [32.8–40.6] | 42.7 [38.9–46.4] |
| Woman | 58.2 [56.0–60.4] | 57.9 [54.2–61.6] | 60.8 [56.9–64.8] | 56.2 [52.4–59.9] |
| In another way | 2.3 [1.7–2.9] | 3.3 [2.1–4.5] | 2.5 [1.3–3.6] | 1.2 [0.5–1.9] |
| Region |  |  |  |  |
| North East | 5.8 [4.8–6.9] | 5.6 [3.7–7.5] | 6.2 [4.2–8.1] | 5.8 [4.1–7.5] |
| North West | 14.7 [13.1–16.4] | 14.7 [11.9–17.5] | 14.6 [11.8–17.5] | 14.9 [12.2–17.6] |
| Yorkshire and the Humber | 11.5 [10.1–12.9] | 11.1 [8.7–13.4] | 13.7 [10.9–16.4] | 10.0 [7.8–12.2] |
| East Midlands | 9.9 [8.6–11.2] | 10.4 [8.1–12.7] | 9.6 [7.3–12.0] | 9.5 [7.3–11.8] |
| West Midlands | 11.0 [9.6–12.4] | 13.1 [10.6–15.6] | 8.0 [5.8–10.1] | 11.4 [9.0–13.8] |
| East of England | 11.1 [9.7–12.5] | 11.1 [8.9–13.3] | 12.0 [9.3–14.7] | 10.4 [8.1–12.7] |
| London | 11.4 [10.0–12.7] | 11.7 [9.4–14.0] | 9.7 [7.4–12.0] | 12.5 [10.1–14.9] |
| South East | 14.1 [12.5–15.7] | 12.0 [9.6–14.4] | 17.4 [14.2–20.7] | 13.3 [10.8–15.9] |
| South West | 10.5 [9.2–11.8] | 10.2 [8.0–12.4] | 8.8 [6.5–11.1] | 12.2 [9.8–14.7] |
| Past-month psychological distress<sup>1</sup> |  |  |  |  |
| None/low | 23.8 [21.3–26.4] | 29.3 [24.7–33.9] | 21.8 [17.2–26.4] | 19.7 [15.7–23.7] |
| Moderate | 35.9 [33.0–38.8] | 36.9 [32.0–41.9] | 38.3 [32.8–43.8] | 32.8 [28.0–37.6] |
| Severe | 40.2 [37.3–43.2] | 33.7 [28.9–38.5] | 39.9 [34.4–45.4] | 47.5 [42.4–52.5] |

Data are presented as weighted column percentages with 95% confidence intervals, unless otherwise specified.
<sup>1</sup> Data on psychological distress were only collected in selected waves (January 2022-June 2023, January-March 2024, and January-February 2025); data shown are valid percentages based on participants not in work due to long-term illness or disability surveyed in these waves (all working-age adults *n*=1,212; never smokers *n*=446; former smokers *n*=349; current smokers *n*=417).

Among those not in work due to long-term illness or disability surveyed in waves that assessed past-month psychological distress, the proportion reporting severe distress was higher among current smokers (47.5%) than former (39.9%) or never smokers (33.7%; **Table 2**). Overall, the proportion who reported severe distress was higher among those aged 18-34 (51.9% [45.1-58.8%]) compared with those aged ≥35 (36.9% [33.7-40.2%]).

## Discussion

This study highlights the extent to which smoking is associated with health-related economic inactivity. Consistent with official data from other national surveys conducted by the ONS,^1,3,4^ we observed a substantial increase in the proportion of working-age adults in England out of work due to long-term illness or disability, with rates more than doubling between 2013 and 2025 – particularly since the start of the COVID-19 pandemic. However, our estimate of the number of working-age adults this represents (1.9 million, as of February 2025) was somewhat lower than the ONS data suggest (2.8 million).^6^ The prevalence of health-related economic inactivity was consistently highest – and absolute increases were largest – among current smokers, which meant the absolute disparity between current smokers and former and never smokers widened over time. By February 2025, one in nine working-age adults in England who smoked was not in work due to long-term illness or disability – approximately 750,000 people – of whom around a quarter were under the age of 35. This figure was up from approximately 390,000 in 2013, despite a decline in smoking prevalence across the period.

Differences by smoking status were more pronounced in older age groups, consistent with evidence that the health impacts of smoking typically become more evident in mid- and later life.^29,30^ In addition, in line with evidence showing benefits of smoking cessation for health and wellbeing,^29,30^ former smokers were less likely than current smokers to be economically inactive due to ill health. We observed a clear dose-response relationship among former smokers, with health-related economic inactivity most prevalent among recent quitters and decreasing with longer durations of abstinence. This suggests that while quitting smoking is beneficial, it does not immediately reverse the health consequences of long-term smoking. However, rates of inactivity were lower among those who had quit for longer periods, emphasising the link between sustained abstinence and improved health outcomes.^30^ These findings suggest that people transitioning away from smoking – particularly those managing chronic health conditions – may benefit from additional support to quit smoking and stay quit to improve their long-term employment prospects.

The sociodemographic profile of people affected by health-related economic inactivity was broadly similar by smoking status. Those who currently smoked were slightly younger than former and never smokers, suggesting that smoking may be contributing to earlier disengagement from the workforce. Given the long-term nature of the health problems associated with smoking, this could leave people at risk of perpetual exclusion from economic opportunities. However, we note that across all smoking statuses, a substantial proportion of those not in work due to long-term illness or disability were aged under 35. Among this younger cohort, psychological distress may be an important factor. Previous studies have documented a sharp rise in distress among young adults in recent years,^19^ and in our analysis of survey waves that included data on past-month psychological distress, half of adults aged under 35 who were not in work due to long-term illness or disability reported experiencing severe distress (a higher rate than among those aged 35 or older). Levels of distress were also notably higher among current smokers than never smokers. These findings highlight the significant mental health burden among economically inactive young adults and people who smoke. The extent to which this a cause or consequence of economic inactivity is unclear and warrants further investigation.

These results provide valuable evidence for economic modelling aimed at assessing the broader economic impact of smoking. Economic models that estimate the cost of smoking-related health issues – such as lost productivity, premature mortality, and healthcare expenditures – rely on high-quality, up-to-date data such as those provided in this study. By quantifying the extent of smoking-related economic inactivity, this study offers key inputs that can inform public health policies and tobacco control strategies. Specifically, the data underscore the need for continued investment in population-level tobacco control interventions (e.g., mass media campaigns) and individual-level support for smoking cessation (e.g., local stop smoking services) as part of broader efforts to reduce long-term sickness-related unemployment and improve overall workforce participation.

Strengths of this study include the large, representative sample and monthly time series, which support the generalisability of the findings to the adult population in England. A key limitation is that the cross-sectional design limits causal inference. While associations between smoking and inactivity are observed, the direction of causality (e.g., whether smoking leads to inactivity or inactivity prompts smoking) cannot be definitively established. However, there is a vast literature showing that smoking is causally linked to a range of serious diseases, causing long-term ill health and disability.^31^ In addition, both the dose-response association we observed between duration of abstinence and health-related economic inactivity among former smokers, and the fact we observed greater differences by smoking status among older age groups (who would have longer smoking histories), suggests a causal relationship. Data were not collected on the specific reason for health-related economic inactivity, so we were unable to explore the contribution of different smoking-associated diseases or mental health concerns to workforce dropout. While findings are most directly relevant to the UK context, similar trends may be observable in other high-income countries with comparable smoking prevalence, healthcare access, and labour market conditions, warranting further international comparison.

In conclusion, current smoking is strongly associated with health-related economic inactivity, and this disparity has widened over time in absolute terms. Efforts to reduce smoking prevalence may contribute to tackling rising inactivity and improving labour market participation.

## Data Availability

Data are available on Open Science Framework (https://osf.io/nygzr/), with age provided in bands to preserve participant anonymity.

## Declarations

### Ethics approval

Ethical approval for the STS was granted originally by the UCL Ethics Committee (ID 0498/001). Participants provide informed consent to take part in the study, and all methods are carried out in accordance with relevant regulations. The data are not collected by UCL and are anonymised when received by UCL.

