## Supplementary material for "A workforce up in smoke? Examining trends in health-related economic inactivity by smoking status in England, 2013-2025": Table S1

**Table S1.** Model selection: AIC values for models with 3, 4, and 5 knots

|  | **AIC** | | |
| --- | --- | --- | --- |
|  | **3 knots** | **4 knots** | **5 knots** |
| All working age adults | 55115.78 | 55092.77 | 55093.90 |
| By smoking status | 53041.24 | 53020.42 | 53025.04 |
| By duration of abstinence, among former smokers | 10066.29 | 10067.57 | 10072.18 |

AIC, Akaike Information Criterion.

Shaded cells indicate the best fitting model (the model with the lowest AIC or the simplest model within 2 AIC units).

**Table S2.** Sample characteristics

|  | **All working-age adults** | **Never smokers** | **Former smokers** | **Current smokers** |
| --- | --- | --- | --- | --- |
| Unweighted *N* | 173,248 | 107,813 | 31,446 | 33,989 |
| Age (years) |  |  |  |  |
| Mean (SD) | 40.5 (13.4) | 39.9 (13.6) | 44.9 (12.4) | 38.3 (13.1) |
| 18-24 | 15.9 | 17.8 | 6.6 | 18.6 |
| 25-34 | 22.1 | 21.8 | 17.7 | 26.9 |
| 35-44 | 21.0 | 20.8 | 22.4 | 20.4 |
| 45-54 | 22.1 | 21.4 | 26.8 | 19.8 |
| 55-64 | 18.9 | 18.2 | 26.5 | 14.3 |
| Gender |  |  |  |  |
| Man | 49.8 | 47.9 | 51.4 | 54.3 |
| Woman | 49.8 | 51.7 | 48.3 | 45.2 |
| In another way | 0.4 | 0.3 | 0.3 | 0.5 |
| Region |  |  |  |  |
| North East | 5.0 | 4.6 | 5.7 | 5.5 |
| North West | 13.2 | 13.0 | 12.8 | 14.5 |
| Yorkshire and the Humber | 10.2 | 9.9 | 10.3 | 10.7 |
| East Midlands | 8.7 | 8.6 | 8.5 | 9.0 |
| West Midlands | 10.3 | 10.6 | 9.7 | 9.8 |
| East of England | 10.9 | 11.0 | 11.0 | 10.7 |
| London | 16.9 | 18.6 | 13.1 | 15.3 |
| South East | 15.4 | 15.0 | 17.7 | 14.7 |
| South West | 9.4 | 8.7 | 11.2 | 9.9 |

Data are presented as weighted column percentages, unless otherwise specified.

**Table S3.** Modelled estimates of changes in the proportion of working-age adults in England not in work due to long-term illness or disability

|  | **% [95%CI] not in work due to long-term illness or disability^1^** | | | | | | | | | | | | |
| --- | --- | --- | --- | --- | --- | --- | --- | --- | --- | --- | --- | --- | --- |
|  | **March 2013** | **March 2014** | **March 2015** | **March 2016** | **March 2017** | **March 2018** | **March 2019** | **March 2020** | **March 2021** | **March 2022** | **March 2023** | **March 2024** | **February 2025** |
| All working-age adults | 2.5  [2.3–2.7] | 2.7  [2.5–2.9] | 2.9  [2.8–3.0] | 3.1  [2.9–3.2] | 3.2  [3.0–3.3] | 3.2  [3.0–3.3] | 3.2  [3.0–3.3] | 3.2  [3.0–3.3] | 3.3  [3.1–3.5] | 3.6  [3.4–3.8] | 4.1  [3.9–4.2] | 4.7  [4.5–5.0] | 5.5  [5.1–5.9] |
| By smoking status |  |  |  |  |  |  |  |  |  |  |  |  |  |
| Never | 1.4  [1.2–1.6] | 1.6  [1.4–1.7] | 1.8  [1.6–1.9] | 1.9  [1.8–2.0] | 2.0  [1.8–2.2] | 2.0  [1.9–2.1] | 2.0  [1.8–2.1] | 1.9  [1.8–2.1] | 2.0  [1.8–2.2] | 2.2  [2.0–2.3] | 2.4  [2.3–2.6] | 2.8  [2.6–3.1] | 3.3  [2.9–3.7] |
| Former | 2.5  [2.1–3.1] | 2.5  [2.2–2.9] | 2.6  [2.3–2.8] | 2.6  [2.3–2.9] | 2.7  [2.4–3.0] | 2.8  [2.5–3.1] | 3.0  [2.7–3.3] | 3.2  [3.0–3.6] | 3.6  [3.2–3.9] | 4.0  [3.6–4.4] | 4.5  [4.2–4.9] | 5.1  [4.7–5.6] | 5.8  [5.0–6.6] |
| Current | 5.4  [4.7–6.1] | 6.0  [5.5–6.5] | 6.6  [6.2–7.0] | 7.1  [6.6–7.6] | 7.4  [6.9–7.9] | 7.4  [6.9–7.9] | 7.2  [6.8–7.7] | 7.1  [6.6–7.7] | 7.3  [6.7–7.9] | 7.8  [7.2–8.4] | 8.7  [8.1–9.2] | 9.9  [9.1–10.8] | 11.3  [9.9–12.7] |
| By duration of abstinence, among former smokers^2^ |  |  |  |  |  |  |  |  |  |  |  |  |  |
| 1 year | 4.7  [3.4–6.5] | 4.8  [3.7–6.1] | 4.9  [4.0–5.9] | 5.0  [4.3–5.9] | 5.2  [4.5–6.1] | 5.5  [4.7–6.5] | 5.9  [5.0–7.1] | 6.5  [5.5–7.7] | 7.3  [6.3–8.4] | 8.2  [7.2–9.3] | 9.3  [8.1–10.7] | 10.7  [9.0–12.7] | 12.1  [9.6–15.1] |
| 5 years | 3.8  [3.0–4.7] | 3.9  [3.3–4.5] | 3.9  [3.5–4.5] | 4.1  [3.7–4.5] | 4.2  [3.8–4.7] | 4.4  [4.0–5.0] | 4.8  [4.2–5.3] | 5.2  [4.6–5.8] | 5.7  [5.2–6.3] | 6.4  [5.9–7.0] | 7.3  [6.6–7.9] | 8.2  [7.3–9.2] | 9.2  [8.0–10.7] |
| 10 years | 3.0  [2.3–3.8] | 3.0  [2.5–3.7] | 3.1  [2.7–3.6] | 3.2  [2.8–3.6] | 3.3  [2.9–3.7] | 3.5  [3.1–3.9] | 3.7  [3.2–4.2] | 4.0  [3.5–4.5] | 4.4  [3.9–4.9] | 4.9  [4.4–5.3] | 5.4  [4.9–6.0] | 6.1  [5.4–6.9] | 6.8  [5.7–8.0] |
| 20 years | 2.3  [1.7–3.1] | 2.3  [1.8–2.9] | 2.3  [1.9–2.7] | 2.3  [2.0–2.7] | 2.3  [2.0–2.7] | 2.4  [2.0–2.8] | 2.5  [2.1–3.0] | 2.7  [2.3–3.2] | 2.9  [2.6–3.4] | 3.3  [2.9–3.7] | 3.7  [3.2–4.2] | 4.1  [3.5–4.8] | 4.6  [3.8–5.7] |

^1^ Data are weighted estimates of prevalence from logistic regression with survey month modelled non-linearly using restricted cubic splines (**Table S1** for model selection), adjusted for age and gender.

^2^ Modelled estimates are shown for selected durations of abstinence to illustrate differences. Note that the model used to derive these estimates included data from former smokers with any duration of abstinence, not only those abstinent for 1, 5, 10, or 20 years.

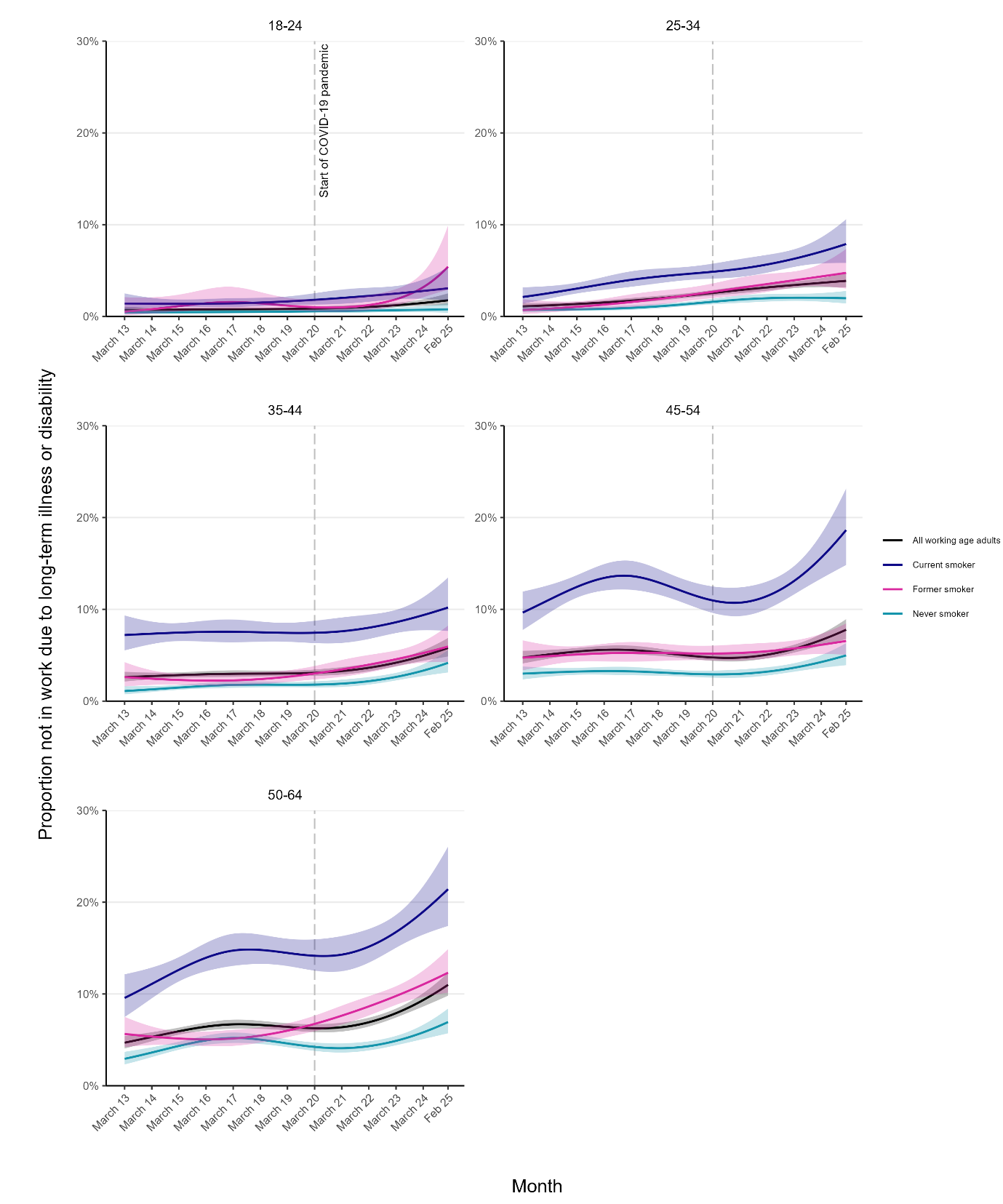

**Figure S1. Age-stratified trends in health-related economic inactivity among working-age adults in England, overall and by smoking status, March 2013 to February 2025.** Lines represent modelled weighted prevalence within each age group by monthly survey wave, modelled non-linearly using restricted cubic splines (four knots), adjusted for age and gender. Shaded bands represent 95% confidence intervals.

**Table S4.** Age-specific modelled estimates of changes in the proportion of working-age adults in England not in work due to long-term illness or disability

|  | **% [95%CI] not in work due to long-term illness or disability^1^** | | **Absolute percentage point change [95% CI]^2^** | **Relative change, prevalence ratio [95% CI]^3^** |
| --- | --- | --- | --- | --- |
|  | **March 2013** | **February 2025** |  |  |
| All working-age adults |  |  |  |  |
| 18-24 | 0.7 [0.5–1.0] | 1.8 [1.2–2.5] | 1.1 [0.4-1.9] | 2.54 [1.41-4.56] |
| 25-34 | 1.1 [0.8–1.5] | 3.9 [3.2–4.7] | 2.8 [1.9-3.7] | 3.56 [2.47-5.13] |
| 35-44 | 2.6 [2.1–3.2] | 5.8 [4.9–6.9] | 3.2 [2.0-4.4] | 2.22 [1.68-2.92] |
| 45-54 | 4.7 [4.1–5.5] | 7.8 [6.8–8.9] | 3.0 [1.7-4.4] | 1.64 [1.31-2.05] |
| 55-64 | 4.7 [4.1–5.4] | 11.0 [9.8–12.3] | 6.3 [4.9-7.9] | 2.34 [1.92-2.85] |
| Never smokers |  |  |  |  |
| 18-24 | 0.5 [0.2–0.9] | 0.8 [0.4–1.4] | 0.3 [0.0-0.9] | 1.70 [0.61-4.34] |
| 25-34 | 0.7 [0.4–1.1] | 2.0 [1.4–2.8] | 1.3 [0.5-2.1] | 2.78 [1.56-5.50] |
| 35-44 | 1.1 [0.7–1.6] | 4.2 [3.1–5.6] | 3.1 [1.7-4.4] | 3.82 [2.16-6.55] |
| 45-54 | 3.0 [2.4–3.8] | 5.0 [3.9–6.3] | 2.0 [0.5-3.6] | 1.67 [1.15-2.38] |
| 55-64 | 2.9 [2.3–3.7] | 6.9 [5.7–8.4] | 4.0 [2.5-5.6] | 2.36 [1.74-3.22] |
| Former smokers |  |  |  |  |
| 18-24 | 0.5 [0.1–2.1] | 5.4 [2.9–9.9] | 4.9 [1.8-8.9] | 10.4 [2.74-134] |
| 25-34 | 0.7 [0.3–1.9] | 4.7 [3.0–7.3] | 4.1 [1.8-6.6] | 6.88 [2.32-32.4] |
| 35-44 | 2.6 [1.6–4.2] | 5.9 [4.3–8.2] | 3.3 [0.8-5.7] | 2.27 [1.23-4.24] |
| 45-54 | 4.7 [3.4–6.6] | 6.6 [5.1–8.5] | 1.8 [0.0-4.2] | 1.39 [0.91-2.21] |
| 55-64 | 5.6 [4.2–7.5] | 12.3 [10.1–14.9] | 6.7 [3.7-9.7] | 2.18 [1.53-3.13] |
| Current smokers |  |  |  |  |
| 18-24 | 1.4 [0.8–2.5] | 3.1 [1.7–5.4] | 1.7 [0.0-3.9] | 2.18 [0.91-5.14] |
| 25-34 | 2.1 [1.4–3.2] | 7.9 [5.8–10.6] | 5.8 [3.3-8.7] | 3.73 [2.15-6.19] |
| 35-44 | 7.2 [5.5–9.3] | 10.2 [7.6–13.5] | 3.0 [0.0-6.9] | 1.42 [0.94-2.18] |
| 45-54 | 9.6 [7.8–11.9] | 18.6 [14.8–23.2] | 9.0 [4.0-13.4] | 1.93 [1.35-2.63] |
| 55-64 | 9.6 [7.5–12.1] | 21.4 [17.4–26.1] | 11.9 [7.0-17.0] | 2.24 [1.62-3.14] |

CI, confidence interval.

^1^ Data are weighted estimates of prevalence in the first and last months in the study period, from logistic regression with survey month modelled non-linearly using restricted cubic splines (four knots), adjusted for age and gender.

^2^ Absolute percentage point change calculated as prevalence in February 2025 minus prevalence in March 2013 with 95% CIs calculated using bootstrapping (1,000 replications).

^3^ Prevalence ratio calculated as prevalence in February 2025 divided by prevalence in March 2013 with 95% CIs calculated using bootstrapping (1,000 replications).
